# Deep Learning for Caries Detection using Optical Coherence Tomography

**DOI:** 10.1101/2021.05.04.21256502

**Authors:** Yu-Ping Huang, Shyh-Yuan Lee

**Affiliations:** Department of Dentistry, National Yang-Ming University; Department of Stomatology, Taipei Veterans General Hospital

**Keywords:** Oral diagnosis, Caries detection, diagnosis, prevention, Digital imaging, radiology, Artificial Intelligence, Deep Learning, Machine Learning, Computer vision, Convolutional Neural networks, Optical coherence tomography, Micro-computed tomography

## Abstract

Early detection of dental caries has been one of the most predominant topics studied over the last few decades. Conventional examination through visual-tactile inspection and radiography can be inaccurate and destructive to the tooth structure. The development of optical coherence tomography (OCT) has given dentistry an alternative diagnostic technique, which has been proven by numerous studies, that it has better sensitivity, specificity, and non-invasive characteristics. The growing popularity of artificial intelligence (AI) also contributes to a more efficient and effective way of image-based detection and decision-making. However, previous studies, which have attempted to employ AI for caries assessment, did not incorporate high-quality ground truth data. Therefore, this study aims to bypass this issue and highlights the importance of high-quality data. A two-phase study was carried out to explore different methods for caries detection. Initially, the comparison of caries detection based on OCT and apical radiography by 5 experienced clinicians was conducted. Then, five convolutional neural networks (CNNs), including AlexNet, VGG-16, ResNet-152, Xception, and ResNext-101, in the scope of AI were employed to detect caries and compared with the findings of the 5 clinicians. The data was preprocessed and labeled with the ground truth corresponding to microcomputed tomography (micro-CT) with rigorous definition. The weighted Kappa statistics suggested that OCT (*ϰ*= .699, SD = .090) showed a higher accuracy rate than radiography (*ϰ*= .407, SD = .049), and CNNs (*ϰ*= .860, SD = .049) were rated higher than clinicians (*ϰ*= .679, SD = .113), both at a .05 significance level. The best result was carried out by ResNet-152, which demonstrated a high accuracy rate of 95.21% and a sensitivity of 98.85%. These findings illustrate the importance of ground truth data for AI training and the potential of deep CNN algorithms combined with OCT for diagnosing dental caries.

## Introduction

Dental caries affects almost 80 percent of children and 90 percent of adults (Bader et al., 2001), thus an effective modality for the early detection of dental caries is an essential topic in dental research (Zandoná and Zero, 2006). Up till now, the most common approach for identifying caries lesions has been through visual and tactile examinations with the help of radiography and dental explorers (Haak et al., 2002). Inspection with radiographic imaging suggests locations of suspicious lesions, followed by tactile examination helping clinicians access carious lesions directly and more accurately. Unfortunately, there are several drawbacks to this method. Firstly, X-ray imaging provides 2D graphs for 3D tooth structures; this means it makes tracing subtle lesions virtually impossible, especially on overlapping structures, including lesions on occlusal, buccal, and lingual surfaces. Thus, intraoral radiography has a poor diagnostic rate of 21% for enamel lesions and 44% for dentin lesions (Molander et al., 1993). Secondly, the tactile examination is experiential and relies on intuition, meaning that there is no quantifiable standard. Furthermore, several studies have indicated that tactile examination may cause irreversible damage to enamel structures (Ekstrand et al., 1987). Finally, the United Nations Scientific Committee on the Effects of Atomic Radiation (2017) also suggests that excessive radiation exposure on infants, children, and adolescents should be avoided, as it can pose more risk than adulthood exposure (e.g. radiogenic tumor induction, cognitive defects, neuroendocrine abnormalities).

Thus far, there has been an array of advanced imaging devices developed for intraoral caries diagnosis, one of which is the Optical Coherence Tomography (OCT). The OCT is a diagnostic tool that relies on an optical interference mechanism (Fercher et al., 2003). There are several advantages that OCT has over traditional radiography. Firstly, data collected from a single OCT procedure consist of multiple scans, in sections, that can be constructed to create a 3D model without interference from adjacent structures, e.g. the alveolar bone surrounding a tooth. Secondly, the high-contrast resolution of OCT scans allows different tissue densities to be distinguished; hence, the possibility of tracing the caries progression is raised. Lastly, since OCT uses low-coherence broadband light instead of tactile movements and X-rays, the risks of tissue damage and radiation exposure are alleviated. Studies have increasingly shown their preference for OCT scans for intraoral caries diagnosis, due to its non-invasive nature and higher sensitivity, compared to radiographs and visual-tactile inspection (Luong et al., 2020; Schneider et al., 2020).

The rapid growth of artificial intelligence (AI), especially convolutional neural network (CNN) in the scope of deep machine learning, reveals an exceptionally suitable method for analyzing and classifying images (Goodfellow et al., 2016). A recent study has also pointed out that these novel algorithms can bring a cost-effective solution to disease detection (Schwendicke et al., 2020). Therefore, it has become increasingly popular in medical analysis. For example, it was implemented in the classification of diabetic retinopathy, as well as, in the detection of cancerous tumor cells (Litjens et al., 2017). These applications have helped CNNs develop a reputation for high recognition rates, accuracy, and efficiency.

As an increasing amount of innovative AI research has been published, a popular saying, “garbage in, garbage out,” has emerged alerting researchers to pay attention to the quality of data and the reliability of the methodology (Rockall, 2020). Recent studies that have attempted to apply CNN algorithms for caries detection have faced limitations. The limitations include labeling carious or non-carious teeth based solely on radiography, as seen in studies done by Srivastava et al. (2017) and Lee et al. (2018), while radiography has been proved with a poor diagnostic rate. Salehi et al. (2019), on the other hand, performed experiments using OCT images without clarifying and addressing how lesions were defined and labeled. Additionally, all above-mentioned studies solely emphasized the detection of caries without considering the “depths” of the lesions. When in fact, effective clinical management is dependent on the awareness of lesion depth (Bader et al., 2001). Lesions limited to the enamel layer require no treatment or merely plaque control; In contrast, operative treatments are required when demoralization and cavities develop in the middle or inner third of the dentin (Ekstrand et al., 2001). Hence, it is vital that further differentiation between lesion seriousness needs to be addressed (Frencken et al., 2012).

Overall, past studies that have attempted to apply CNN algorithms to caries detection have lacked validity and reliability on methodology. The impracticable classification also limited the potential for clinical decision-making. Therefore the objectives of this study were, within a valid and reliable method, to establish a ground truth micro-CT images to provide quantitative research on diagnostic rates of radiography and OCT, and compare the diagnostic outcomes between experienced clinicians and the CNN models.

## Materials and Method

### Imaging Techniques

A self-developed swept-source OCT (SS-OCT) was used in this study. The system operated at a center wavelength of 1310 nm with an average power of 40 mW, a scan rate of 50kHz, and a frame rate of 160 fps. Each frame contained 200 scanning lines, and the resolution of each 2D image was 250 × 1024 *px*. Further detailed illustration, settings, and mechanisms are shown in Appendix A.

The periapical films of the teeth samples were acquired with ScanX Intraoral Phosphor Plates (Air Techniques Inc., USA) and PY-70M Intraoral Imaging Systems (POYE Inc., Taiwan), operating with 70 *kV p* tube potential, 10 *mA* tube current, and 0.7 x 0.7 mm focus point. The films were processed with ScanX-Duo D1000F Digital Radiography System, exported in .*tiff* format with the resolution of 652 × 801 *px*.

Micro-CT is a device used for assessing dental caries by analyzing the mineral contents of teeth through non-destructive characteristics (Swain and Xue, 2009). The device is currently considered one of the best for assessment, even though it still relies on clinician judgement and verification. Therefore, Micro-CT was chosen to be the judgment basis in this study, as it can reach a deeper depth than OCT scans and present high-resolution data. The Micro-CT used in this study was High-Resolution U-CT (Milabs, Netherlands), operating with 50 *kV* tube voltage, 0.48 *mA* tube current, and 10 *µm* resolution.

### Convolutional Neural Network (CNN)

CNN is well-known for its powerful capability for image processing. Instead of conventional architectures that connect all the perceptrons costing tremendous computational time and power, CNN adopts a more efficient method by recognizing hierarchical patterns. Detailed CNN basics can be further observed in Appendix B.

Throughout this study, using CNN algorithms for the detection and classification of caries were our main technique of interest. To gauge its effectiveness, the results were compared to experienced clinician detection and classification of caries. This allows us to determine the feasibility of using CNNs as assistance in differential diagnoses. A high recognition rate, accuracy, and efficiency were hypothesized. The CNN models adopted in our study were: AlexNet (Krizhevsky et al., 2012), VGG-16 (Simonyan and Zisserman, 2014), ResNet-152 (He et al., 2016), Xception (Chollet, 2016), and ResNeXt-101 (Xie et al., 2017), which are the classic and best models for image recognition over the recent years. The CNN model architectures adopted in our study remained the same as their original published versions. Additionally, the PyTorch framework (Paszke et al., 2019), based on Python programming language, was used in this study as the implementation of deep learning models.

### Data Collection

The study was approved by the Institutional Review Board of National Yang-Ming University (YM109025E). One hundred teeth with different levels of caries lesion were collected and immersed in 5% sodium hypochlorite (NaOCl) for a week for disinfection and then stored in distilled water. Fractured teeth, previously restored teeth, and teeth with obvious cavities that can be visually differentiated, were excluded, leaving 63 teeth for further study. For each tooth, we took one micro-CT scan as the reference, one periapical film and one scan of OCT for our research. The periapical films were clipped into a unified size (591 × 421 *px*); on the other hand, the OCT data was converted via our self-developed software to 200 sections .*bmp* images, with a unified size (420 × 209 *px*).

All the experimental data required classification. The micro-CT data and periapical films were coded according to a simplified version, shown in Table 1, of the widely-used criterion for radiographic examination formulated by Ekstrand et al. (1997) (see Appendix C). Our self-developed criterion used for classifying the OCT images is also described in Table 1, which was based on the criterion developed by Shimada et al. (2010) (see Appendix D). The implementation of rigorous labelling assured the codes of all experimental data to be mutually exclusive and independent. Supplemental pictures are shown in Fig. 1.

**Table 1:** Criteria of carious status classification in radiography and OCT

| Code | Criteria |
| --- | --- |
| Radiography: Periapical film and Micro-CT |  |
| 0 | No radiolucency visible |
| 1 | Radiolucency visible in the enamel, but not involved in the dentine |
| 2 | Radiolucency visible in the outer, middle, and pulpal $\frac{1}{3}$ of the dentine |
| OCT |  |
| 0 | No caries. Obtained OCT signal was the same level and shape as that of normal enamel and loss of enamel surface did not occur. |
| 1 | Superficial demineralization of enamel or enamel breakdown due to the caries. OCT signal intensity was enhanced within the enamel thickness, whereas continuity of enamel surface may be disconnected at the occlusal fissure or not. |
| 2 | Dentine caries. An intensified OCT signal was obtained beyond the EDJ, with or without loss of enamel surface (cavitation). |

**Figure 1:**
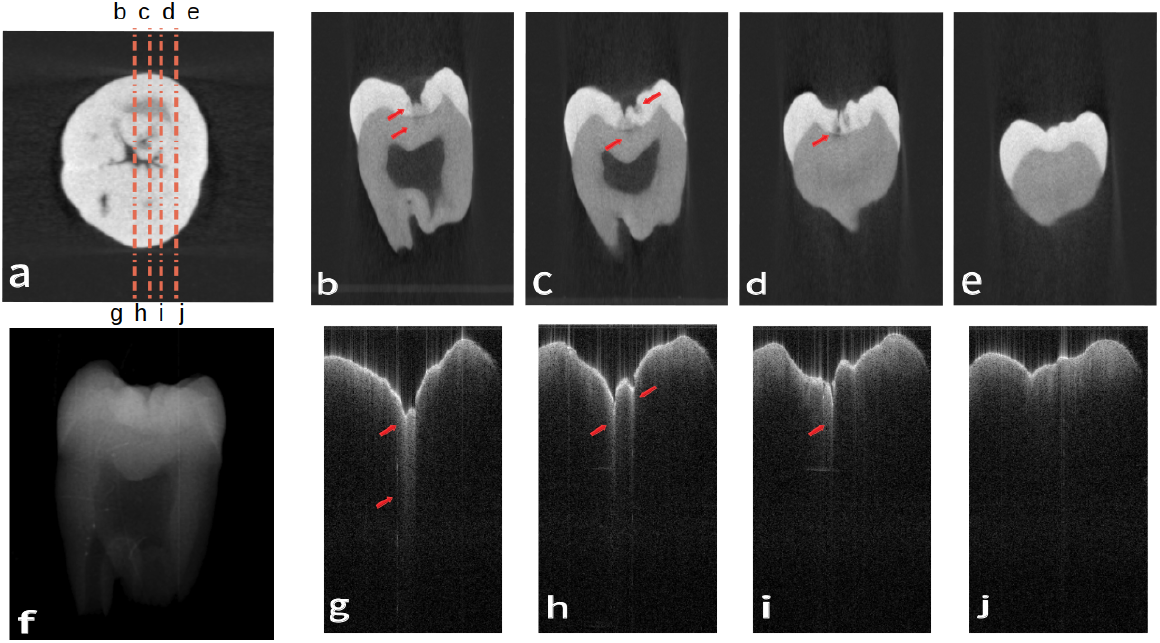
Images of three different image data. (a) is the micro-CT data of a sample tooth in coronal view. (b)(c)(d)(e) correspond to four different segment as illustrated in (a). (f) is the periapical film of the tooth. (g)(h)(i)(j) are the OCT counterparts to (b)(c)(d)(e) of (a). Carious lesions are indicated at the points of red arrows, where discontinuous enamel surface and intensified OCT signals. With the corresponding criteria, (b)(c)(g)(h) are coded as ⟨2⟩ for dentine caries with an intensified OCT signal beyond the EDJ. (d)(f)(i) are coded as ⟨1⟩ because of the superficial demineralization and enamel breakdown. And (e)(j) are coded as ⟨0⟩, represent intact tooth sections without any caries.

### Procedures

A two-phase study was conducted to explore different methods of carious examination. The first section aimed to verify the accuracy and effectiveness of OCT scans. To prove the hypothesis that the diagnostic capabilities of OCT images were better than periapical films. In the second section, the study shifted our objective to the comparison of interpreters, clinicians and CNN algorithms, to see if CNN algorithms were comparable with clinicians or even better in diagnosis. A supplemental flowchart is depicted in Appendix E.

#### Phase 1: Comparison of Diagnostic Capability of OCT versus Radiography

We took periapical films and OCT data of the sample teeth and asked five well-educated clinicians to examine the images. The clinicians were asked to make judgments of carious status individually for each tooth based on both types of data, following the criteria shown in Table 1. Afterward, their answers were scored and evaluated with the corresponding ground truth obtained by Micro-CT. The weighted Kappa coefficient was then calculated. Since this measurement took the possibility of the accidental match and the correlation in lesion extent into account, it is more reliable and convincing than pure accuracy (Ben-David, 2008; Chmura Kraemer et al., 2002).

The participants in this study were five resident doctors in the Dental Department of Taipei Veterans General Hospital. Before participating, they were informed that our study was interested in determining whether OCT or Radiography is better at caries detection; they were, however, not told what types of results were expected. The unit of comparison was a “tooth” to ensure some homogeneity between the two different data types.

Given the inherent disadvantages of radiographs compared to multi-sectional OCT data, if the clinicians asked for more images for confirmation, we provide additional shifted images with the commonly-used tube-shift technique (Seiler et al., 2018). Additionally, if there were any discrepancies between clinicians’ diagnoses of radiographs, a discussion was allowed, and a correction based on the mutual agreement was noted. To avoid any possible misunderstanding, the answer given under such circumstances was recorded independently and denoted as “CR” (Consensus on Radiography). This methodology was designed to encourage the best radiography diagnosis. However, clinicians were not allowed to discuss their diagnosis when using the OCT scans and were not given any supplemental data. Hence, if the results still show that OCT has superiority over radiography, it may indicate that OCT is the more effective method of caries detection.

#### Phase 2: Comparison of Diagnostic Ability of clinicians versus CNNs

The OCT data was once again given to five clinicians and processed using the five above-mentioned CNN models to see if there was a significant difference in their ability to detect caries.

Since the CNNs required a large database, the unit of the sample was changed from “tooth” to “image”. Therefore, a total of 748 cross-sectional 2D images were extracted from 63 OCT tooth data, which were then labeled into three distinct groups according to Table 1: 470 images with ⟨0⟩, 174 images with ⟨1⟩, and 104 images with the code ⟨2⟩. Images were divided into 599 (80%) training data and 149 (20%) testing data with opposite intentions. Training data was responsible for optimizing the parameters in the CNN models, while testing data evaluated the models. After the CNNs finished their training session, the test data was sent to both trained CNN models and clinicians for diagnoses. The obtained results were statistically analyzed and compared.

Similarly to phase 1 of the study, we offered clinicians the best possible accommodations. If there were any discrepancies amongst clinician diagnosis, discussion and correction based on the mutual agreement were permitted. The answers given under such circumstances were once again recorded independently and denoted as “CC” (Consensus of Clinicians). Hence, if the results still favored CNNs over clinicians, it may indicate that CNNs are more effective in caries detection compared to clinicians.

Finally, we further evaluated the performances of the result from the best CNN model. Considering that related researches were conducted in binary classification, we reformulated our best result into the corresponding form. The purpose was to assess the clinical feasibility of the outcome. The statistical measurement consisted of accuracy, sensitivity, specificity, positive predictive value (PPV), and negative predictive value (NPV), all of which were considered essential indexes in medical screening.

## Results

The diagnostic capabilities of OCT and periapical radiography, from phase 1 of the study, were compared in Table 2. Each individual weighted Kappa coefficient was computed, followed by the calculation of mean value, standard deviation, variance, together they summarize the average performance and data distribution. The mean of the previously defined “CR” periapical radiography result was also calculated, allowing for comparisons to be drawn. Then the *p*-value was calculated, based on the Mann-Whitney U test between the Radiography and OCT groups, to examine the significance of differences between the clinicians and CNNs groups.

**Table 2:**
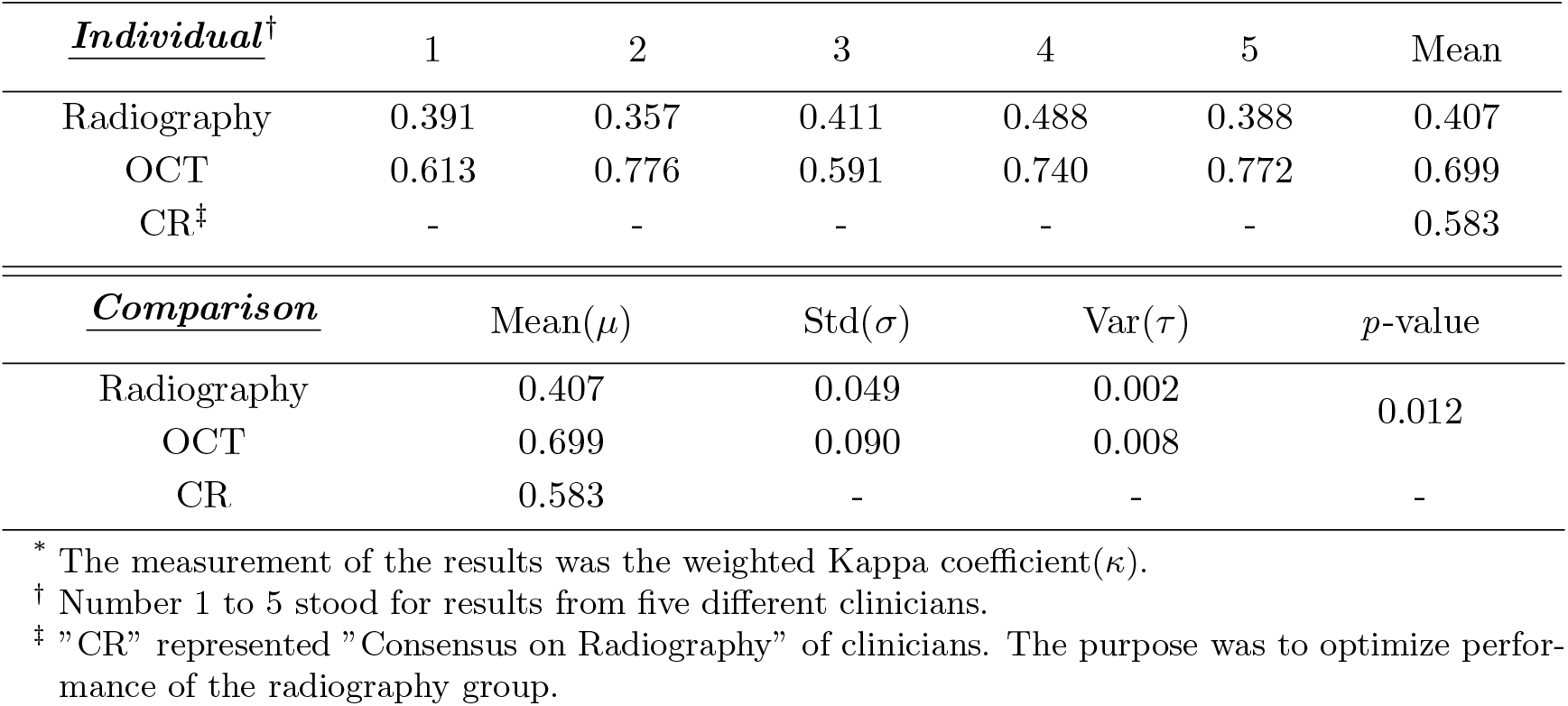
Diagnostic capability* of periapical radiography and OCT

The results show that clinicians scored a higher Kappa value with OCT (*M* = .699, *SD* = .090) compared to radiography (*M* = .407, *SD* = .049) with a statistically significant difference at the .05 level (*p* = .008). However, the variance of OCT (*τ* = .008) was slightly bigger than that of radiography (*τ* = 0.002). While, the “CR” answer showed a lower Kappa value (*ϰ* = .583) than that of OCT (*ϰ* = .699).

Given the affirmation that OCT had better diagnostic capability compared to periapical radiography, CNN algorithms were prompted to read OCT images in order to compare their diagnostic with clinicians in phase 2. The same statistical analysis was used in phase 2, as in the previous phase, and was tabulated in Table 3.

**Table 3:** Diagnostic ability* of clinicians and CNNs based on OCT

| ***Individual*** | 1 | 2 | 3 | 4 | 5 | Mean($\mu$) |
| --- | --- | --- | --- | --- | --- | --- |
| Clinicians<sup>†</sup> | 0.633 | 0.715 | 0.505 | 0.783 | 0.760 | 0.679 |
| CNNs<sup>‡</sup> | 0.891 | 0.852 | 0.855 | 0.915 | 0.786 | 0.860 |
| CC<sup>§</sup> | - | - | - | - | - | 0.668 |

| ***Comparison*** | Mean($\mu$) | Std($\sigma$) | Var($\tau$) | $p$-value |
| --- | --- | --- | --- | --- |
| Clinicians | 0.679 | 0.113 | 0.013 | 0.012 |
| CNNs | 0.860 | 0.049 | 0.002 |  |
| CC | 0.748 | - | - | - |

| ***(Best)*** | Accuracy(%) | Sensitivity (%) | Specificity(%) | PPV (%) | NPV (%) |
| --- | --- | --- | --- | --- | --- |
| ResNet-152 | 95.21 | 98.85 | 89.83 | 93.48 | 98.15 |

\* The measurement of the results was the weighted Kappa coefficient ( $\kappa$ ).
<sup>†</sup> Number 1 to 5 stood for results from five different clinicians.
<sup>‡</sup> Number 1 to 5 stood for five CNN models in the following order: AlexNet, VGG-16, ResNet-152, Xception, and ResNext-101.
<sup>§</sup> "CC" represented "Consensus of Clinicians". The purpose was to maximize the performance of the clinicians group.

As shown in Table 3, CNNs (*M* = .860, *SD* = .049) outperformed clinicians (*M* = .679, *SD* = .113) on average when comparing the Kappa value. The variance of CNNs (*τ* = .002) was also much smaller than that of clinicians (*τ* = .013). The difference between the two groups was statistically significant at the .05 level (*p* = .012). However, the mean values in the “CC” group (*M* = .748) were still lower than those in CNNs (*M* = .860). A visual comparison box plot can also be found in Fig. 2, illustrating the Kappa value in each group.

**Figure 2:**
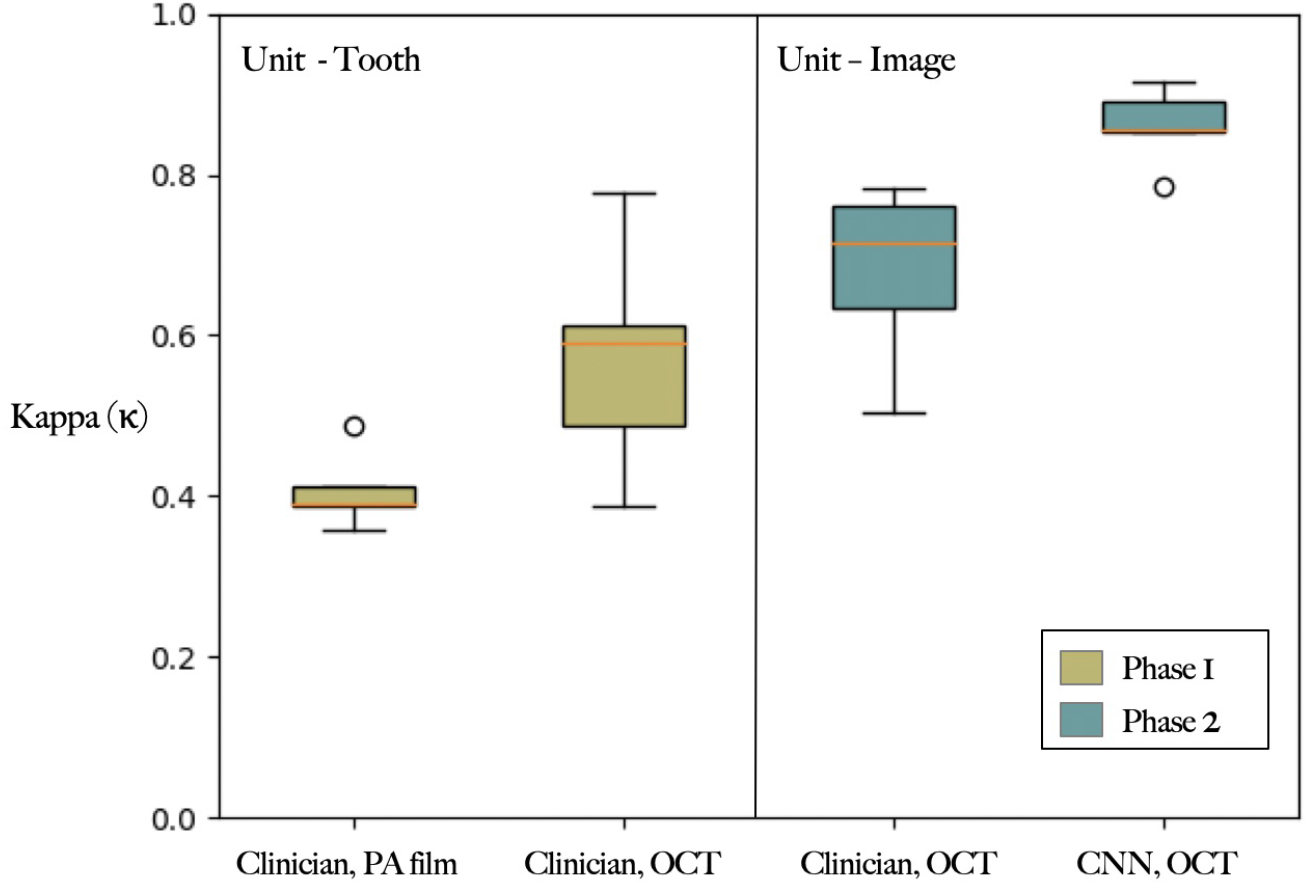
Comparison of diagnostic capability of Radiograph versus OCT and diagnostic ability of clinician and CNN. Note that the calculation in different sections was based on different objects – teeth or images.

The best experimental model, ResNet-152, was further evaluated, as shown in Table 3. The model differentiated caries lesions on OCT images with an accuracy of 95.21%. More specifically, the sensitivity was 98.85%, the specificity was 89.83%, and the PPV and NPV were 93.48% and 98.15%, respectively.

## Discussion

The majority of clinicians currently still rely on tactile examination and radiography to make dental caries diagnostics. However, this approach has proved to have low accuracy rates and is highly dependent on the doctor’s training background, work experience, and concentration during the examination. The advanced imaging tool, OCT, working in tandem with the rise of AI, has provided a potentially better solution for caries detection. Increasing studies have attempted to use AI, OCT, or a combination of both to enhance caries detection. However, most of them lacked a valid and reliable methodology, and failed to achieve high-quality classification.

In order to develop a better detection technique, a reliable methodology based on high-quality data needs first to be developed. In our two-phase study, the well-defined criteria of caries classification and a reliable judgment basis, the Micro-CT, was implemented as the reference for scoring. The results indicated that clinicians could recognize carious patterns via OCT images more accurately than via periapical radiographs. Even the accuracy of clinicians in identifying radiographs is enhanced after discussion, it is still not comparable to that of OCT scans, let alone this is unrealistic in chairside to have a group of experienced clinicians discuss a single scan. However, the slightly higher variance in the OCT group revealed more uncertainty regarding OCT than radiography interpretation; this could be due to experience with radiographs. Furthermore, CNNs yielded excellent repeatability of results among OCT and Micro-CT more so than clinicians, which indicates that CNNs can detect and classify carious lesions more precisely and more consistently. The findings overall suggest that OCT is more effective than radiography in caries detection, and CNNs performed higher accuracy and consistency than clinicians on these tasks.

Our results were generally in line with previous studies (Srivastava et al., 2017; Lee et al., 2018; Salehi et al., 2019), however, are more well-rounded and address aspects that were overlooked in previous studies. Our study focused on the establishment of a detailed classification method that is more valid and reliable.

The validity of data is a central issue within the scope of deep machine learning. Without strict examination, the results at most correspond with existing labels but often have little application to real-world situations. If the labels are wrong, then even if the results completely matched with the labels, the results would have no value. Compared to previous studies, our data was compared to a more reliable device, the Micro-CT, which allowed for better diagnosis and also reached deeper depths of teeth. Additionally, all the definitions of classification were clearly delivered and evaluated through content analysis, hence, proves that this study has a higher criterion-related validity.

Secondly, while there is a general agreement that the seriousness of carious lesions plays a critical role in diagnosis and should be involved when considering the appropriate treatment plan, precise classification in previous studies were barely emphasized. In this study, we separated lesions of different degrees of demineralization and activity, particularly enamel and dentin lesions, to enhance treatment decision-making processes.

Moreover, while the architectural innovations of CNN have grown rapidly in recent years, related studies only adopted a singular model. This study adopted five different models, from the classical to the most innovative ones. AlexNet was first used in 2012 and a breakthrough in CNN, followed by VGGNet, which inherited the concept of AlexNet and deepened the model for better outcomes. However, it was not until ResNet that successfully solved the problem of gradient degradation by residual learning. ResNet, which allowed for a more profound model architecture to emerge, demonstrated extraordinary accuracy on image recognition. Soon afterwards, Xception and ResNext were developed mainly to enhance model performance with minimal efficiency sacrifice. Among all, the best model for our specific task was ResNet-152. According to our results, the high sensitivity (98.85%) and NPV (98.15%) were the most critical indicators in clinical trials, as they ensure a low false-negative rate and imply that few caries lesions would be missed. We highly recommend ResNet to future OCT readings and related applications. Overall, this study aimed to address the weakness overlooked by previous studies to verify their results and provide much more enhanced results.

Although this study has yielded significant results, the potential limitations should be noted; the most prominent limitation in this study was the manual verification process. The labels had to be manually marked by researchers, even using Micro-CT as a correspondence truth, where human errors are inevitable. A possible solution to alleviate this error in future research is through the use of image registration technique, which is commonly used in brain mapping but rarely validated in dental radiography. This would also allow the depths, extents, and boundaries of caries lesions to be automated corresponded. Large amounts of direct labels would be provided without manual labor, allowing for more detailed and precise detection study results. While this study has its limitations, it can still serve as a basis for further studies in related topics.

To the best of our knowledge, this study is the first to use Micro-CT as a solid reference, develop a three-tier classification and adopt five different CNN models. With the rigorously defined experiments accompanied by comprehensive statistical analysis, the validity and compatibility of our results are more integrative and reliable. Our extensive methodology and experimental results are of great interest for further scientific research and clinical application, respectively. Hopefully, in the future, more AI-based clinical studies can include strict and reliable methodology.

## Conclusion

A clinically applicable automatic diagnostic methodology on caries detection was posed in this study. The results indicate that CNNs are better than clinicians at distinguishing pathological tooth structures using SS-OCT images. The findings lead us to believe that CNNs, with appropriate imaging techniques, have a large potential and practicality especially when implemented in clinics to provide patients with more adequate diagnoses.

## Data Availability

The data that support the findings of this study are available on request from the corresponding author (Lee, SY). The data are not publicly available because they contain information that could compromise research participant privacy/consent.

## Author Contribution

Yu-Ping Huang, contributed to conception, design, data acquisition, analysis, and interpretation, drafted and critically revised the manuscript; Shyh-Yuan Lee, contributed to conception, design, and critically revised the manuscript. All authors gave final approval and agreed to be accountable for all aspects of the work.

## Acknowledgment

This research was partially supported by the Ministry of Science and Technology (MOST 108-2813-C-010-040-B) and the Department of Health, Taipei City Government (10901-62-017). The quality of this experiment was greatly enhanced by the assistance of Dr. Lyu Dong-Yuan, who assisted in setting and adjusting the OCT instrument. We would also like to thank Dr. Chen Li-Fen, Inst. of Brain Science, for their invaluable consultations and advice on deep learning. Additionally, the authors would like to thank all other colleagues who contributed to this study. All authors gave their final approval and agree to be accountable for all aspects of the work.

## Appendices

### A Mechanism of Our SS-OCT

The mechanism of our swept-source OCT (SS-OCT) system is illustrated in Appendix Fig. 3. A beam of light initially generated from the swept-source laser is separated into two parts right after going into the coupler and circulator. While one of them enters the sample arm, the other goes into the reference arm, both of which are then reflected. After reflection, the separated light beams merge back together. Optical path differences are created due to the different characteristics of the surfaces, resulting in distinctive interference patterns.

**Figure 3:**
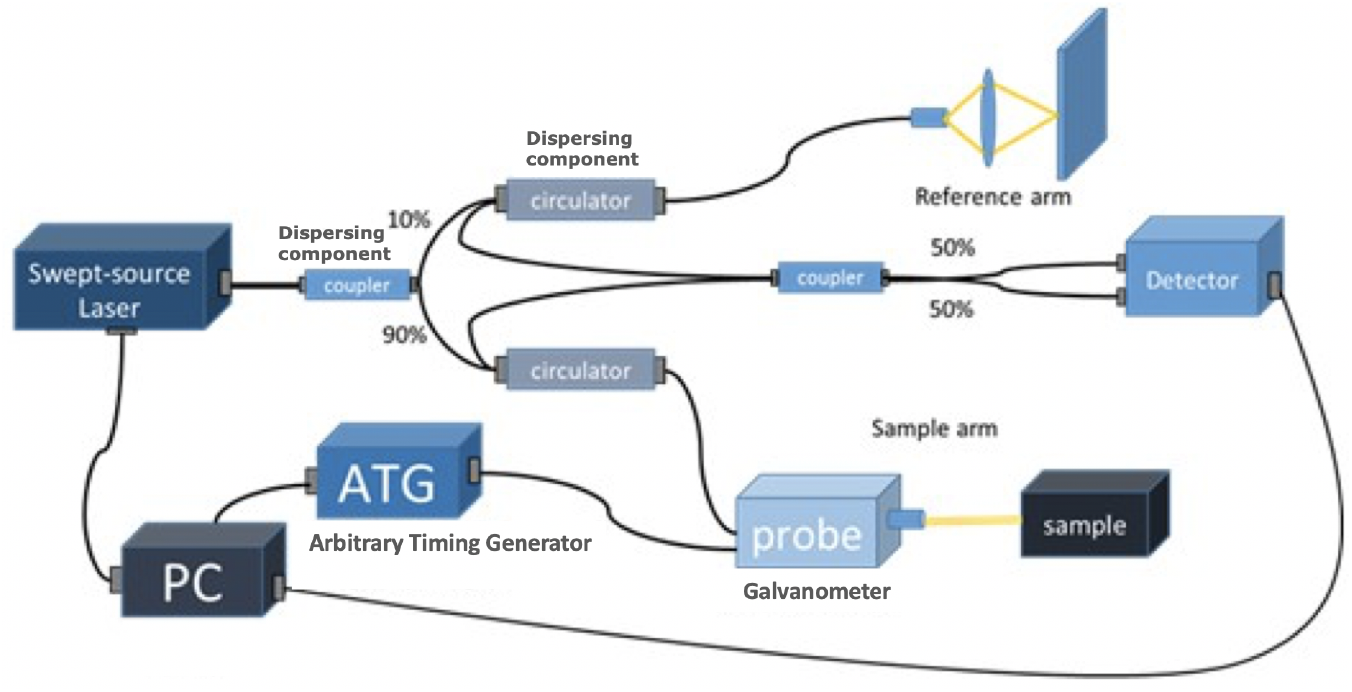
Setup of our SS-OCT scanning device

### B Introduction of Convolutional Neural Network

The basic structure was first carried out by LeCun et al. (1998) as shown in Appendix Fig. 4, which consists of an input and output layer with several convolution and pooling layers in between.

**Figure 4:**
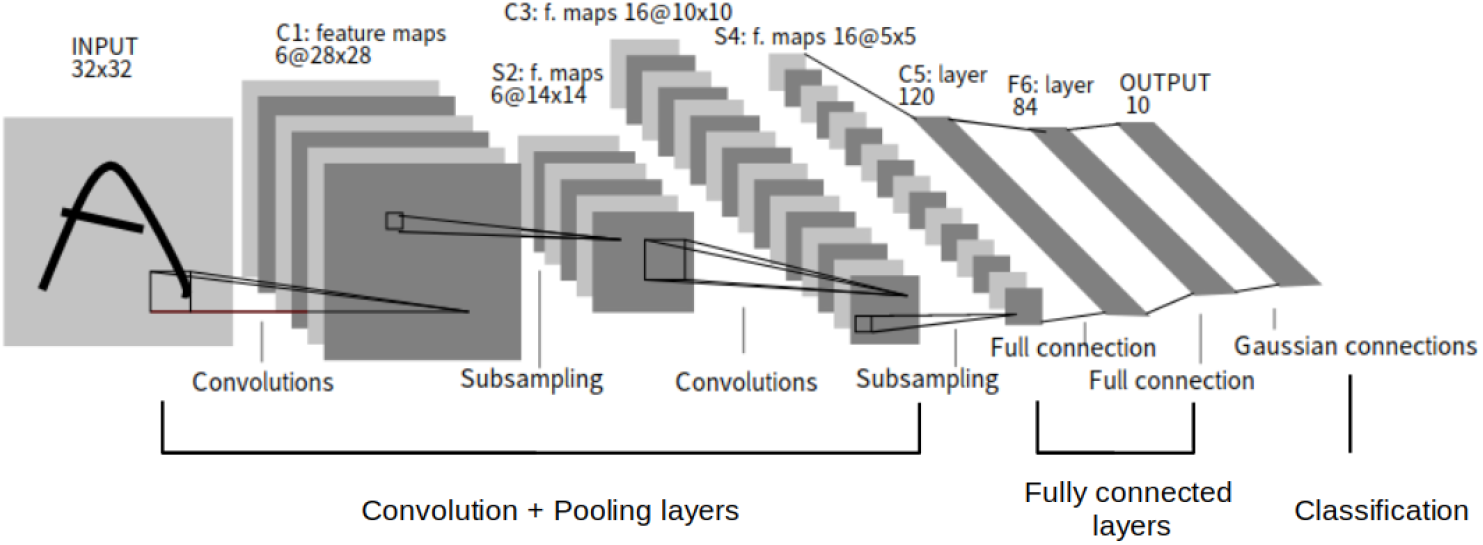
Illustration of a CNN model architecture

#### (a) convolution

A convolution procedure is illustrated in Appendix Fig. 5. In the context of image processing, images (denoted *A*) are modeled as real matrices by its pixel value. Convolution serves as a binary operation among matrices of different shapes. For 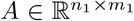 and 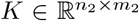, *A* * *K* would yield another matrix 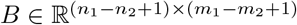,

**Figure 5:**
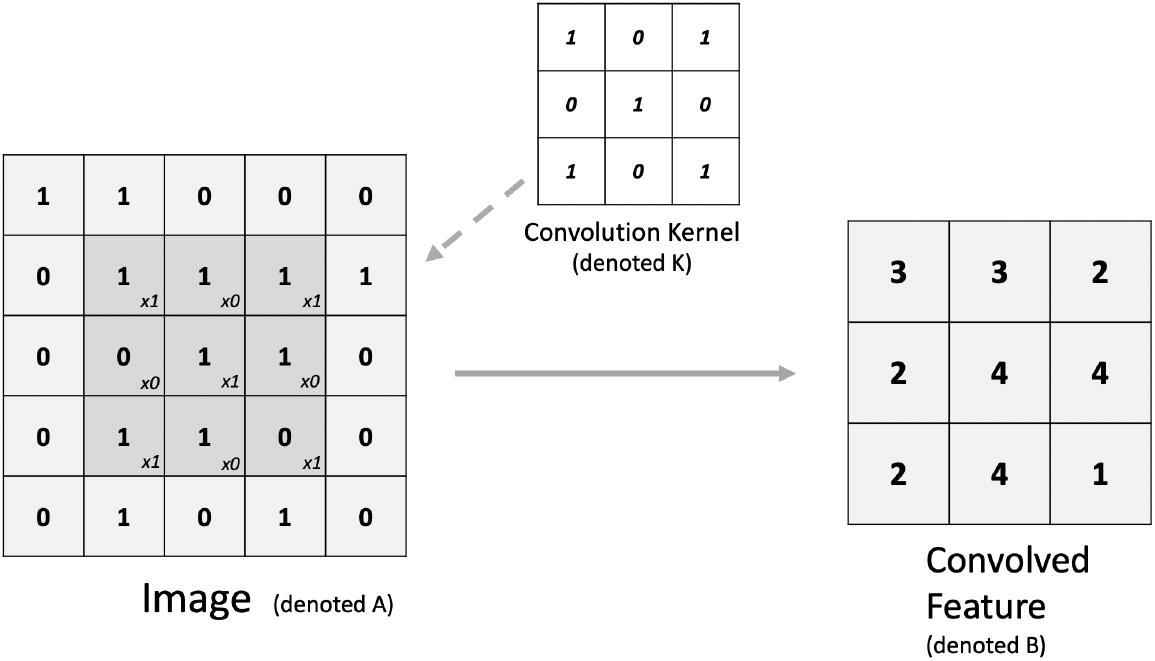
Basic structure and function of a convolution layer

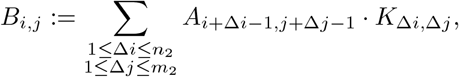

where *K* is usually much smaller than *A* and referred to as the *kernel*. Convolutions with small kernels are usually for purposes of local feature extraction such as detecting edges, impulses or noise in images. Usually one needs to design an appropriate kernel, but the presence of convolution layers is to find the appropriate kernel via parameter optimization. We can view the convolution as a bi-linear operator

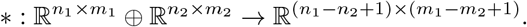

Sometimes, there can be multiple channels for both images and kernels. This motivates one to have the *multi-channel convolution* by linearly extending the original one. For 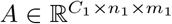 and 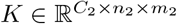 where *C*_1_, *C*_2_ are the numbers of channels for *A, K* respectively. The resulting *B* = *A* * *K* would be simply

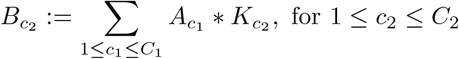

#### (b) pooling

The pooling technique decreases computational resources without losing much information. The most common methods are max-pooling and average-pooling as shown in figure 6. In a max-pooling layer of stride *s* and width *w*, a image *A* ∈ ℝ^*n*×*m*^ is mapped to *B*1,

**Figure 6:**
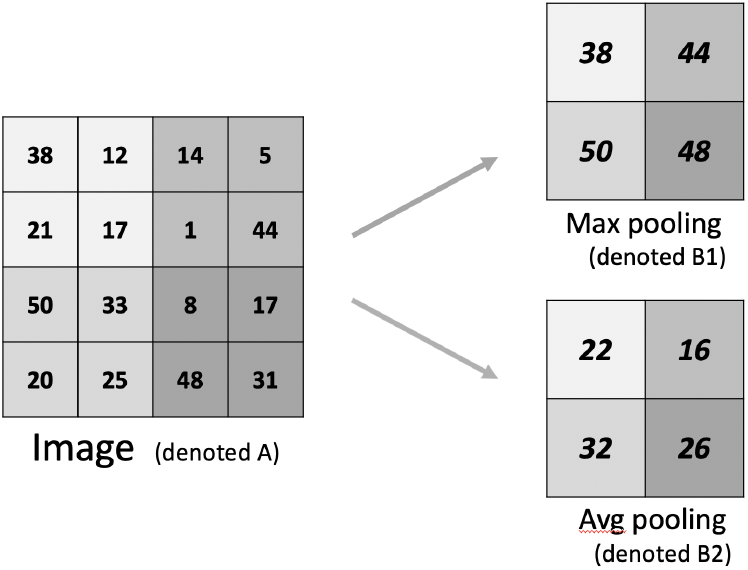
Demonstration of max-pooling and average-pooling techniques

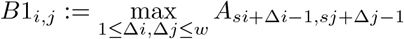

On the other hand, for an average-pooling case, we have *B*2,

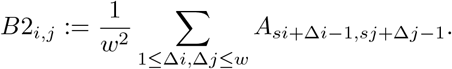

Interleaving convolution and pooling layers, concatenated by a fully connected layer at the end, would yield a complete CNN model which mimics an image processing pipeline. A CNN model takes in an image then output a probabilistic vector. Finally, it uses the well-known back-propagation algorithm to optimize its parameters in each layers until the model generalize well to data.

The CNN models adopted in our study were: AlexNet, VGG-16, ResNet-152, Xception, and ResNeXt-101. All are thought of as the best models in different periods. More specific architecture details should be found in the original papers (as in the references).

### C Caries Classification in Radiographic Examination (Ekstrand et al., 1997)

| Code | Criteria |
| --- | --- |
| 0 | No radiolucency visible |
| 1 | Radiolucency visible in the enamel |
| 2 | Radiolucency visible in the dentine but restricted to the outer $\frac{1}{3}$ of the dentine |
| 3 | Radiolucency extending to the middle $\frac{1}{3}$ of the dentine |
| 4 | Radiolucency in the pulpal $\frac{1}{3}$ of the dentine |

### D Caries Classification of OCT Examination (Shimada et al., 2010)

| Code | Criteria |
| --- | --- |
| 0 | No caries. Obtained OCT signal was the same level and shape as that of normal enamel and loss of enamel surface (cavitation) did not occur. |
| 1 | Superficial demineralization of enamel. OCT signal intensity was enhanced within the enamel thickness but loss of enamel surface (cavitation) did not occur. |
| 2 | Enamel breakdown due to the caries. Continuity of enamel surface is disconnected at the occlusal fissure, where OCT signal was intensified but limited to the enamel thickness. |
| 3 | Dentine caries. An intensified OCT signal was obtained beyond the EDJ, with or without loss of enamel surface (cavitation). |

### E Overall Procedure of This Study

The workflow of this study is depicted in Appendix Fig. 7. The main objects of comparison are painted: the beige sets and the green sets represent the objects in the first and second parts, respectively, while the blue box (Micro-CT) is recognized as the reference of the ground truth.

**Figure 7:**
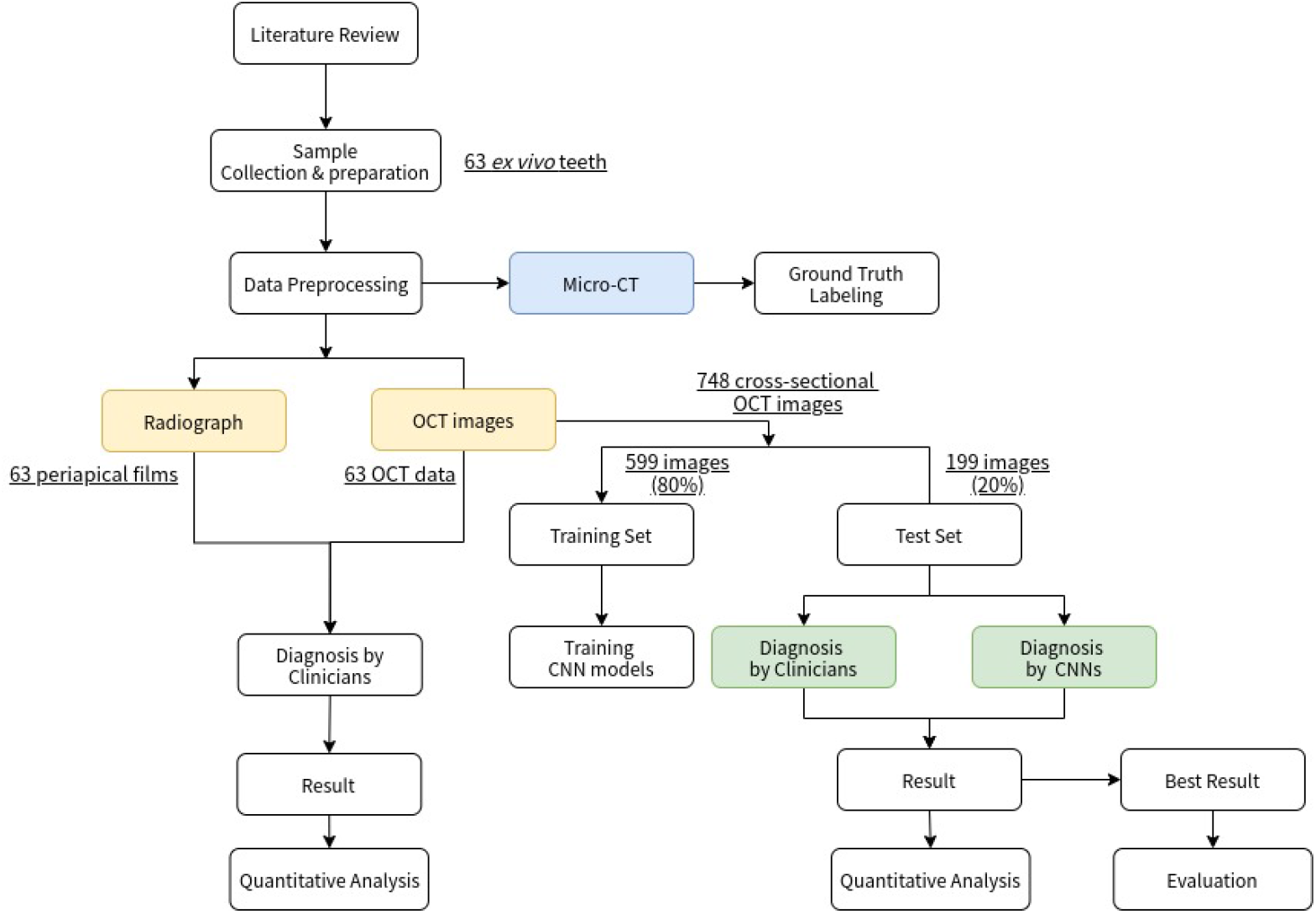
Illustration of the procedure in this study.

